# Diagnostic accuracy of Panbio™ rapid antigen tests on oropharyngeal swabs for detection of SARS-CoV-2

**DOI:** 10.1101/2021.01.30.21250314

**Authors:** Marie Thérèse Ngo Nsoga, Ilona Kronig, Francisco Javier Perez Rodriguez, Pascale Sattonnet-Roche, Diogo Da Silva, Javan Helbling, Jilian A. Sacks, Margaretha de Vos, Erik Boehm, Angèle Gayet-Ageron, Alice Berger, Frédérique Jacquerioz-Bausch, François Chappuis, Laurent Kaiser, Manuel Schibler, Adriana Renzoni, Isabella Eckerle

## Abstract

**Background:** Antigen-detecting rapid diagnostic tests (Ag-RDTs) for the detection of SARS-CoV-2 offer new opportunities for testing in the context of the COVID-19 pandemic. Nasopharyngeal swabs (NPS) are the reference sample type, but oropharyngeal swabs (OPS) may be a more acceptable sample type in some patients.

**Methods:** We conducted a prospective study in a single screening center to assess the diagnostic performance of the Panbio™ COVID-19 Ag Rapid Test (Abbott) on OPS compared with reverse-transcription quantitative PCR (RT-qPCR) using NPS.

**Results:** 402 outpatients were enrolled in a COVID-19 screening center, of whom 168 (41.8%) had a positive RT-qPCR test. The oropharyngeal Ag-RDT sensitivity compared to nasopharyngeal RT-qPCR was 81% (95%CI: 74.2-86.6). Two false positives were noted out of the 234 RT-qPCR negative individuals, which resulted in a specificity of 99.1% (95%CI: 96.9-99.9) for the Ag-RDT.

For cycle threshold values ≤ 26.7 (≥ 1E6 SARS-CoV-2 genomes copies/mL, a presumed cut-off for infectious virus), 96.3% sensitivity (95%CI: 90.7-99.0%) was obtained with the Ag-RDT using OPS.

**Interpretation:** Based on our findings, the diagnostic performance of the Panbio^™^ Covid-19 RDT with OPS samples meet the criteria required by the WHO for Ag-RDTs (sensitivity≥80% and specificity ≥97%).

## Introduction

The SARS-CoV-2 pandemic has killed millions of people worldwide(1). Large scale testing allows for identification and isolation of infected individuals, and quarantining contacts, thus limiting community transmission. Currently, SARS-CoV-2 RT-qPCR performed on nasopharyngeal swabs (NPS) is the gold-standard diagnostic test. While displaying excellent sensitivity and specificity, RT-qPCR is costly, subject to reagent and material shortages during pandemics, and requires experienced personnel and complex infrastructure. Antigen rapid diagnostic tests (Ag-RDTs) are easy to use, more affordable, decentralizable, and provide quick results; offering an attractive alternative to RT-qPCR during pandemics. Their drawbacks are mainly reduced sensitivity relative to RT-qPCR.

The World Health Organization (WHO) considers a sensitivity ≥80% and a specificity ≥97% as acceptable performance for SARS-CoV-2 Ag-RDTs(2). Currently, only validations of Ag-RDTs performed with NPS have shown satisfactory results(3– 12), and no studies have evaluated Ag-RDTs using oropharyngeal swabs (OPS). OPS sampling could be a useful alternative to NPS sampling, as seen with RT-qPCR tests(13,14). Here we describe a prospective study comparing the diagnostic performances of an Ag-RDT using OPS with RT-qPCR using NPS for detection of SARS-CoV-2.

## Methods

### Ethics

The study was approved by the cantonal ethics committee (Commission Cantonale d’Ethique de la Recherche, CCER, Geneva, Nr. 2020-02323). All enrolled patients provided written informed consent form.

### Setting, study design and participants

The study took place from November 3 to 19, 2020, at an outpatient SARS-CoV-2 screening site at the Geneva University Hospitals. The majority of patients had symptoms compatible with SARS-CoV-2 infection, and a small proportion were asymptomatic contacts. All participants were ≥16 years old.

### Sampling procedure

Participants were swabbed twice: one NPS performed by a nurse at the screening site, for the reference RT-qPCR; and an OPS done by an experienced doctor, using a tongue depressor in a well-lit environment with an emphasis on consistent technique, for the Ag-RDT.

A pilot study tested 28 RT-qPCR-positive individuals without ensuring the back of the oropharynx was reached, yielding only 11 Ag-RDT positives (Supplemental table 1). Therefore, patients were only included if the posterior wall of the oropharynx could be reached.

**Table 1:**
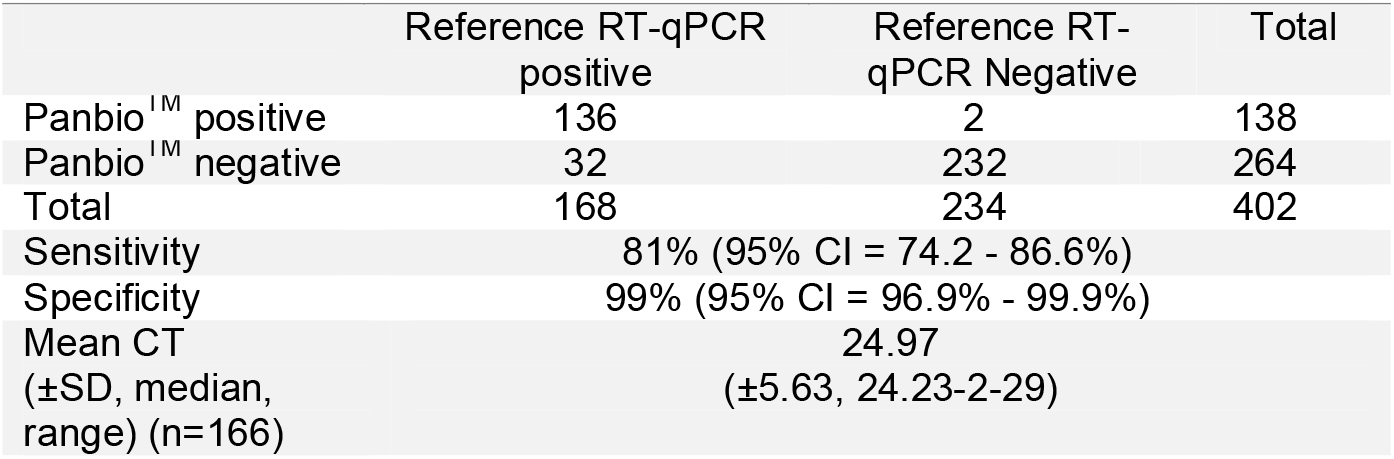
Diagnostic performance of the Panbio™ rapid antigen test in oropharyngeal specimens

### Data collection

The clinical data collected for each patient was: duration of any symptoms when samples were collected, potential close contact with a positive person within 14 days, symptoms (rhinorrhea, odynophagia, myalgia, chills, dry vs productive cough, hemoptysis, fever, anosmia, ageusia, gastrointestinal symptoms, asthenia, dyspnea, chest pain and headache), and comorbidities (hypertension, cardiovascular disease, chronic lung disease, diabetes, chronic renal failure, active cancer, severe immunosuppression, pregnancy and obesity (BMI> 40 kg/m2)).

### Ag-RDT procedure

Aside from the sample type, the Panbio™ (Abbott) Ag-RDT device was run and read by a biologist according to the manufacturer’s protocol. Equivocal results were read by a second healthcare worker.

### SARS-CoV-2 detection by RT-qPCR

All NPS samples were analyzed using the Cobas^®^ SARS-CoV-2 RT-PCR assay on the 6800 system (Roche), targeting ORF1 and the E-gene. Cycle-threshold (Ct) values for the E-gene were converted into viral load (VL) with the following formula: log_10_ SARS-CoV-2 copies/mL=(Ct-44.5)/-3.3372.

### Statistical methods

Ag-RDT sensitivity and specificity was determined relative to RT-PCR. With a positivity rate of 37.5%, and an Ag-RDT sensitivity/specificity of 85%/95%, a sample size of 400 could determine sensitivity and specificity with confidence intervals (CI) of 79.3-90.7% and 92.3-97.7%, respectively. Fischer’s exact test was used to compare Ag-RDT sensitivity by Ct values (above/below 26.7). All analyses were performed using STATA version intercooled 16 (Stata Corp., College Station, TX, USA). Statistical significance was defined as p<0.05 (two-sided).

## Results

During the study period, 402 participants were included. Eight patients were excluded either because the throat was insufficiently accessible or because consent was withdrawn. The participants’ socio-demographic characteristics are summarized in supplemental table 2. 168 participants (41.8%) were RT-qPCR-positive with a mean Ct value of 24.97 (SD ±5.63, 3.3E6 SARS-CoV-2 copies/mL equivalent) for 166 RT-PCR analyses. Two specimens, positive for the ORF1 target at a high CT values but negative for the E-gene, were interpreted as positive in the analysis for sensitivity and specificity but excluded from Figure 1; both were Ag-RDT negative.

**Figure 1.**
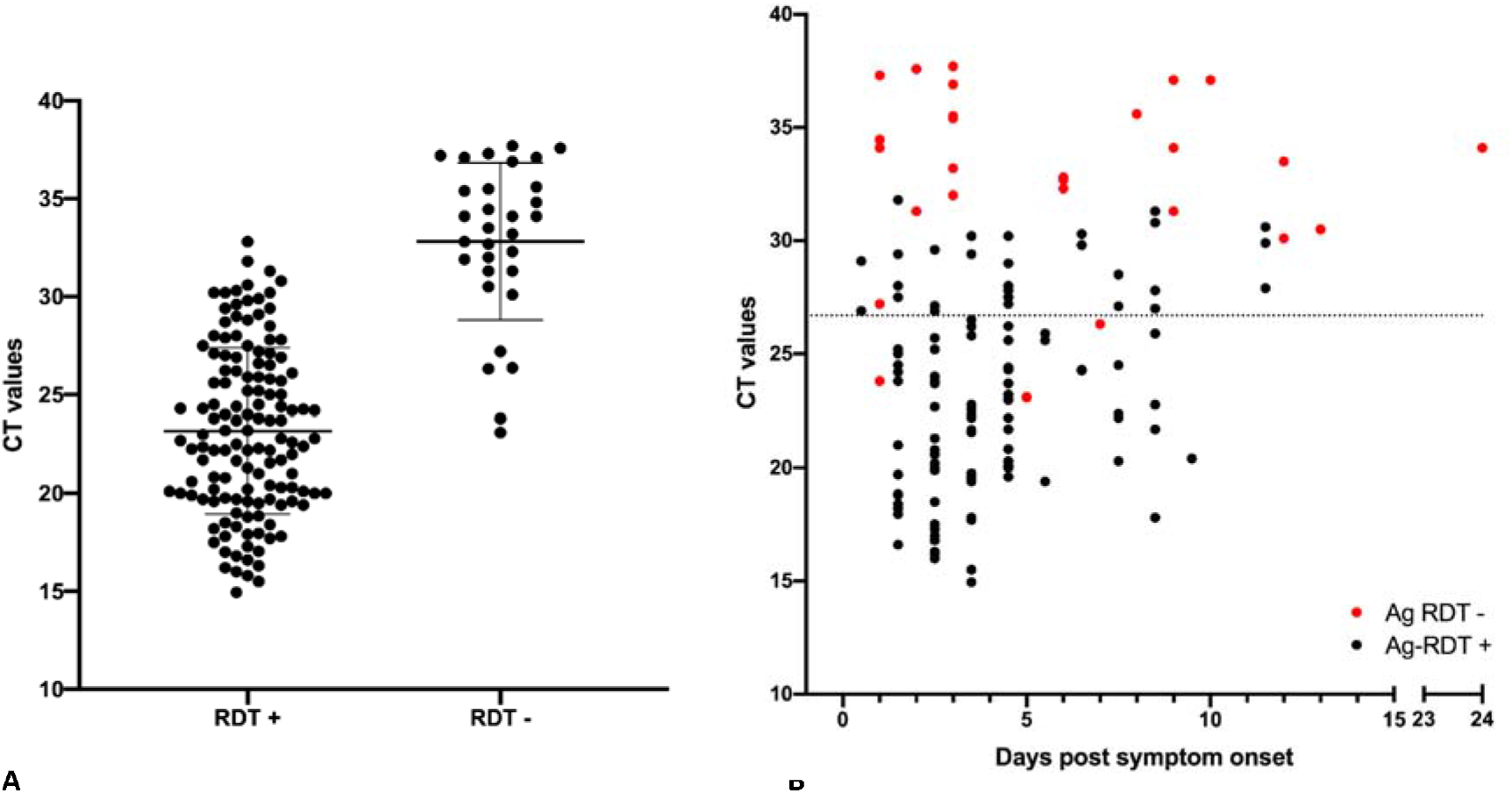
SARS-CoV-2 detection by Panbio™ antigen rapid test using OPS compared to the reference RT-qPCR detection method using NPS. A. Ct values, viral load and Ag-RDT results for 166 RT-PCR-positive individuals. Horizontal bars represent median and standard deviation. Dotted line: Ct value of 26.7 or 1E6 SARS-CoV-2 RNA copy numbers/mL. Note: Two samples were excluded because of low viral load (positive signal in ORF1 assay but negative signal in E-gene target, thus excluded from the graph. Both samples gave a negative RDT result). **B**. Ct values, viral load, days post symptom onset and Ag-RDT results for 139 patients for which information on day of symptom onset was available. Dotted line: Ct value of 26.7 or 1E6 SARS-CoV-2 RNA copy numbers/mL.

All RT-qPCR-positive participants were symptomatic. Compared to RT-qPCR, the sensitivity of the Ag-RDT was 81% (95%CI: 74.2-86.6). Two Ag-RDT false-positives were observed, thus the specificity was 99.1% (95%CI: 96.9-99.9) (**Table 1**).

The sensitivity of the test for Ct values ≤26.7 (equivalent to ≥1E6 SARS-CoV-2 copies/mL) was 96.3% (90.7-99.0%). Of the OPS samples from RT-PCR-positive individuals, mean Ct value for Ag-RDT-positive samples was 23.17 while the mean Ct value for Ag-RDT-negative samples was 32.82, equivalent to 1.1E7 and 1.3E4 SARS-CoV2 copies/mL, respectively (**Figure 1**).

Ag-RDTs have shown higher sensitivity in individuals with lower Ct values/higher VL, and in the first days post onset of symptoms (DPOS)(3). As false-negative Ag-RDT results correlate with low VLs, we expected higher numbers of false negative results in samples collected later after the onset of symptoms.

For patients presenting within 0-4 DPOS, the sensitivity was 86.1% (n=101; 95%CI: 77.8-92.2). For those presenting within 5-7 and 8-11 DPOS, it was 73.7% (n= 19; 95%CI: 48.8-90.9) and 70.6% (n=17; 95%CI: 44.0-89.7), respectively. Sensitivities in the presence of fever or chills; fever and cough; fever and anosmia or fever and cough; and non-specific symptoms, were: 87.5% (n=80; 95%CI: 78.2-93.8), 92.3% (n= 39; 95%CI: 79.1-98.4), 92.5% (n=53; 95%CI: 81.8-97.9), and 84.0% (n=25; 95%CI: 63.9-95.5), respectively.

## Discussion

There are over 10 clinical studies (8 preprints, 2 published) evaluating the performance of the Panbio™ Ag-RDT(3–12) using only manufacturer recommended NPS. Those studies, with over 6000 subjects, have reported sensitivity and specificity ranges of 71.4%-91.7% and 94.9%-100%, respectively. Considering only Ct values <30 yielded test sensitivities from 87.7% to 97.8%(3,6–9). Similarly, samples from <5 DPOS yielded a sensitivity between 77.2 and 94.8%(3,5,6,8,9).

For some patients in whom NPS sampling is not feasible, OPS could be an attractive alternative, thus OPS sample validation is critical. This is the first publication investigating the diagnostic accuracy of the Panbio™ Ag-RDT using OPS. Our results meet show this off-label use to still meets the WHO targets of ≥80% sensitivity and ≥97% specificity(2). Interestingly, while ensuring high OPS sample quality, we obtained similar results to our previously published NPS evaluation, with no statistical difference in sensitivity and specificity(3).

We previously demonstrated similar clinical and analytical sensitivities between NPS and OPS sampling for SARS-CoV-2 detection using RT-qPCR(14). However, some studies showed reduced(13) sensitivity and lower rates of virus isolation in cell culture for OPS when compared to NPS, suggesting a risk of reduced Ag-RDT sensitivity when using OPS(15).

Our present study shows that despite the use of OPS, contrary to manufacturer recommendations, we obtained highly reliable results. Similar to studies on NPS specimens, the highest sensitivity was seen in the early symptomatic period as well as for patients presenting with high nasopharyngeal VL. Although a few positive samples with lower Ct values were missed, the majority of false-negative samples were from individuals with high Ct values (≥30), corresponding to a low VL below the presumed cut-off for infectious virus (Ct≤26.7 in our hands); these results suggest that these individuals are not likely to be contagious and that these false-negative Ag-RDT results should not result in further transmission.

In conclusion, the use of Ag-RDTs with OPS might prove to be an acceptable alternative to NPS, and could increase test acceptance for selected groups such as children.

## Data Availability

Data will be available upon request to the corresponding authors. Due to ethical consideration, not patient-specific data can be shared.

## Acknowledgements

We thank all nurses and staff at the testing Centre Sectors of our institution as well as the patients for their willingness to participate in the study.

## Supplemental Tables and Figures

**Supplemental Table 1:**
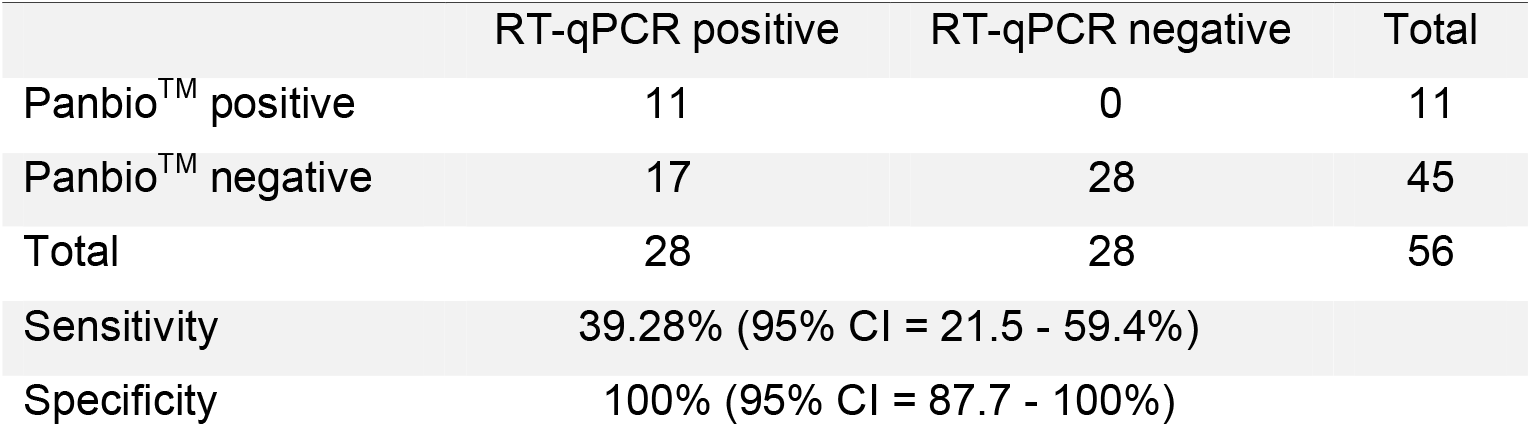
Results of the pilot study

**Supplemental Table 2.** Patient Characteristics

Characteristics
|  |  |
| --- | --- |
| Mean age (±SD, median, range) | 39.9 (±14.5, 38, 16-80) |
| Sex distribution, n (%) |  |
| Women | 224 (55.7) |
| Men | 178 (44.3) |
| Mean days from symptom onset to PCR<br>(SD, Median, range) (n=141) | 4.1 (±3.3, 3, 0-24) |
| Comorbidities among positive PCR, n (%) | 38 (22.6) |
| Number of patients with symptoms, n (%) | 168 (41.8) |
| Symptoms, n (%) (n=168) (descending<br>order in frequencies) |  |
| Asthenia | 101 (60.1) |
| Headaches | 99 (58.9) |
| Myalgia | 81 (48.2) |
| Chills or fever | 80 (47.6) |
| Dry or productive cough | 73 (43.5) |
| Anosmia, ageusia | 71 (42.3) |
| Odynophagia | 68 (40.5) |
| Digestive signs | 38 (22.6) |
| Dyspnea | 7 (4.2) |
| Chest pain | 4 (2.4) |
| Other | 12 (7.1) |
| Had a contact with positive cases within last<br>14 days, n (%) | 87 (51.8) |

